# The Influence of Parkinson’s Disease on Depression Through Structural Equation Modeling

**DOI:** 10.1101/2024.05.29.24308144

**Authors:** Jasmine Moon, Jonah Winninghoff, Mariam Paracha

**Affiliations:** Department of Psychology, University of Rochester, Rochester, NY; Rochester Institute of Technology, Rochester, NY; Gallaudet University, Washington DC; Department of Neurology, University of Rochester Medical Center, Rochester, NY

**Keywords:** Parkinson’s Disease, depression, nonmotor symptoms, structural equation modeling, directed acyclic graph, item response theory, quality of life, apathy

## Abstract

**Objective:** To analyze and compare the magnitudes of nonmotor symptoms in depressed individuals with and without Parkinson’s Disease.

**Methods:** Downloaded in January 2023, symptoms and characteristics of depressed individuals with and without Parkinson’s Disease were extracted from the longitudinal study Fox Insight. The directed acyclic graph theory was used to represent causal relationships and to determine the associations between nonmotor symptoms, depression, and Parkinson’s Disease. The 2-PL and the Rasch models were used to better fit the data and identify the items consistent with the assumptions.

**Results:** *Dropped interests and activities* was the item that was of greatest impact and most characteristic for depressed individuals both with and without PD. *Double vision* was the item of least difficulty while an additional item of significant impact was *Preferred to stay at home*. Education attainment had little to no impact on depression severity in individuals with and without PD.

**Conclusions:** *Dropped interests and activities* was most severe within both populations. Relatedly, an increased average number of leisure hours was associated with greater depression severity. Overall, as an individual’s depression worsens, there is an overall upward trend in the severity of their nonmotor symptoms. We recommend additional analyses on the strength of relationships between these neurologic disorders and nonmotor symptoms.

## INTRODUCTION

Parkinson’s Disease (PD) is a progressive, neurological disease characterized by motor and nonmotor symptoms (NMS) including depression or anxiety, cognitive impairment, and sleep disorder.^1,2^ NMS frequently occur in the earliest stages of PD and may precede the onset of motor disability by over a decade.^3^ In fact, they may have an equal, if not greater, impact on the quality of life (QOL) than motor symptoms,^3,4^ and mood symptoms, and decreased functional ability are key determinants of reduced QOL for individuals with PD (iPD).^5^

Depression affects an estimated 3-4 million iPD, making depression one of the most common NMS, but it is underrecognized and undertreated.^6,7^ The diagnosis of depression in iPD is complicated by the overlap of psychopathological symptoms and the motor features of PD.^4,8,9^ Due to equivocal findings, literature regarding the influence of depression in iPD has been inconclusive.^2^

We have two primary hypotheses. Aligning with findings of similar studies on this topic, we also hypothesize that symptoms of psychopathological impairment related to changes in mood or behaviors will be most severe among individuals with both depression and PD. Relatedly, our second hypothesis is that the presence of PD among individuals, all else being equal, leads to an increase in the severity of depressive symptoms.

Thus, in this paper, the objective is to analyze and compare the magnitudes of NMS in depressed individuals with and without PD using the Item Response Theory (IRT) approach to determine a factor score, which is an estimate of the quantitative value manifested by an individual’s latent traits. The association between specific NMS symptoms and depression in iPD remains unclear so the IRT approach can help identify specific symptoms that differentiate the depressed individuals with PD from those without PD. Previously, a few studies focused on the effect of depressive symptoms on a PD diagnosis but have not concurrently studied these depressive symptoms for both individuals with and without PD. Thus, the second objective of this study is to specify the linear and nonlinear regression models-using the directed acyclic graph (DAG) approach-to compare the individuals with and without PD.

## METHODS AND PROCEDURES

### Data and sample selection

Fox Insight is a longitudinal survey sponsored by the Michael J. Fox Foundation for Parkinson’s Research that accounted for more than 7,000 variables on at least 50,000 adults (Fox Insight, Data Exploration Network). This database collected information about NMS, medical conditions, labor force activity, income, health coverage, and some demographic characteristics of those with and without PD. This survey conducted assessments every three months for over five years. The data used in this study was downloaded in January 2023.

The eligibility criteria for this study were that every respondent was diagnosed with ongoing depression, and they completed all survey items in respect to NMS. The longitudinal dataset showed that the number of respondents over the age of 18 who meet the eligibility criteria plummeted when considering the second survey, so this paper focused on the first respondent-reported values-treating this analysis as cross-sectional. Therefore, the total sample size consisted of 545 respondents. The minimum sample size necessary for the fixed effect linear regression model with R-squared deviation from zero with power equal to 95%, medium effect size (f^2^=0.15), and six independent variables was at least 146; this minimum sample size number was determined using the *G*Power* software, so our sample size was more than sufficient. The Rasch model used in this study required the sample size to be equal to or greater than 150 for consistent estimation while the required sample size for the 2-parameter logistic (2-PL) model fell between 250 and 750, depending on the number of items.^11^

### Directed Acyclic Graph Theory

The DAG theory is used to represent causal relationships **(Figure 1)**. The DAG helps identify the variables that need to have conditions applied and those that can be left unconditioned and is used to determine whether the depressed iPD have a higher factor score, indicating a stronger association with the symptom, than the depressed individuals without PD. The DAG consists of nodes and edges with arrows but does not have a feedback cycle, which is a capture of causal pathways among variables. If the directed edge X→Y, then the X causes Y. Additionally, the definition of confounding bias is more precise in the AG. For example, X←Z→Y may be identified as a confounder while a collider is when X→Z←Y. The mediator is when X→Z→Y. Except for the colliders, confounders and mediators should be conditioned for model specification.^12^

**Figure 1.**
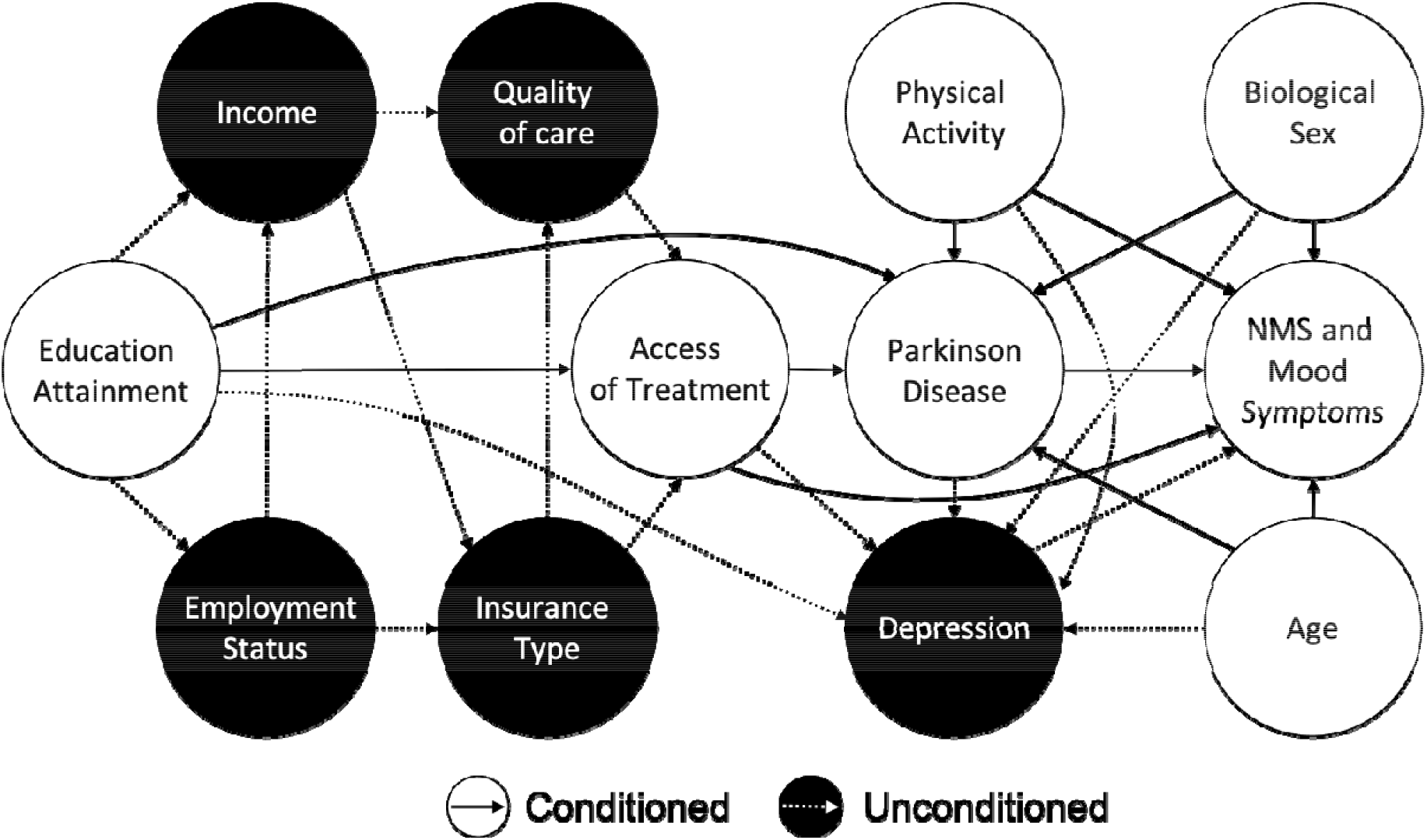
Directed Acyclic Graph of Parkinson’s Disease and NMS.

The DAG represents relationships among variables that encompass NMS. The conditioned paths represent variables being included in the regression model while the unconditioned paths represent variables that are excluded.

Despite the challenges in quantifying access to treatment, there is a likelihood of an association between access to treatment and parkinsonian and depressive symptoms. For example, pharmacogenetic testing is used to identify a specific set of genes that can help determine the appropriate medication and dosage for individuals, minimizing the risk of adverse side effects. The number of unique treatment respondents used is a proxy variable for measuring access to treatment. In this study, we also recognize the diagnosis with PD and the presentation of NMS may coincide with biological sex, physical activity, education, and age.^14-17^

### Latent Variable Analysis

The IRT is a family of mathematical models using the measurable items to analyze the latent variable. In this paper, two mathematical models, Rasch and 2-PL models, were used. The 2-PL model aimed to better fit the data while the Rasch model aimed to identify the items consistent with the IRT assumptions.

As the nature of the Rasch model is an item calibration, this model consists of no more than one parameter that measures the difficulties of items and latent trait of individuals. Each NMS-related item has dichotomous values. The item difficulty corresponds to the point of median probability-indicating the severity of depressive symptoms in terms of 50% of individuals confirming this symptom. Individuals with a lower depression severity are less likely to confirm the more difficult items. This model was used to identify distortions in measurements and correlations among items.

In the 2-PL model, one added parameter is item discrimination-a measure of sensitivity probability when latent traits change. This measure determines how informative each item is and detects the inadequate items when their discrimination score is below zero. For example, when an individual confirms an item that is higher in discrimination, it is distinguishable from the rest of the individuals with a similar latent trait. Furthermore, the 2-PL model is more likely than the Rasch model to be consistent with various indexes of fits.^18^ The reliability of both models depends on the IRT assumptions. Prior to using the 2-PL model, the Rasch model tested all assumptions. These tests utilized an empirical dataset with 95% confidence interval to estimate the position from item characteristics curve.

The 2-PL model was tested for its capability to fit the data using three metrics: root mean square error of approximation (RMSEA), comparative fit index (CFI), and Tucker-Lewis index (TLI). While CFI and TLI are indices used to compare the fit of a hypothesized model to that of a baseline model, RMSEA is used to test how different the hypothesized model is from a perfect model.^19^ Furthermore, the expectation maximization was used to estimate the parameters. The *eRm* and *mirt* softwares in the R program were used to analyze the latent variable.

### Regression Model

At least one item that signals the difference between the groups of depressed individuals with and without PD may be coincidental. Thus, the evaluation of the difference was first modelled with the ordinary least squares (OLS), using the factor score as a dependent variable. The corner solution is a special case variable not resulted by censoring observations. This phenomenon is not an observable issue. Because the problem with the corner solution arises from this analysis, the maximum likelihood was then applied to specify this model called Tobit. Based on the 2-PL model, the lowest factor score among the 54 respondents was -1.25. The Tobit model produced consistent estimates.^20^

This analysis was reported with both standard errors and bootstrapped standard errors. The bootstrapped standard errors were in use to remediate the non-normality issue in residual distribution. This analysis produced these results using *VGAM*, *vglm*, *lmtest*, *sandwich*, and *stats* softwares in the R program. The censReg software was in use to obtain the ATE for the Tobit model.

## RESULTS

### Latent Variable Result

The 2-PL model included a set of NMS items, and the Rasch model showed that these eight items were reliable and consistent **(Table 1)**. Using Principal Component Analysis, 29.12% of explained variance was captured while at least 20% was acceptable.^21^ There was no significant correlation with each item according to the Q_3_-based nonparametric model using the Benjamini-Hochberg procedure. As indicated by the Wald test, none of these items were flagged as differential item functioning (DIF) in biological sex groups, which means the results of these responses were not statistically significant. The indices of fit concluded that the 2-PL model fit well with the dataset (CFI=1; TLI =1.638; RMSEA=0). In the 2-PL model, none of these items exhibited significant distortion of the measurement system as evaluated by infit mean squared statistics. However, as indicated by standardized mean statistics, this model is overfitted.

**Table 1.** Coefficients of 2-PL Model.

| Item description | Discrimination ($a_i$) | Difficulty ($b_i$) |
| --- | --- | --- |
| **Nonmotor** |  |  |
| 1. Hallucinations/delusions | 1.341 | 2.025 |
| 2. Double vision | 0.844 | 2.469 |
| 3. Unpleasant sensations | 0.702 | 0.466 |
| 4. Frightening dreams | 0.777 | 0.640 |
| 5. Excessive sweating | 0.738 | 1.171 |
| **Mood** |  |  |
| 6. Dropped interests and activities | 1.792 | 0.335 |
| 7. Preferred to stay at home | 0.789 | -0.776 |
| 8. Memory difficulties | 0.982 | 0.803 |
| 9. Often feeling helpless | 1.540 | 0.768 |

As indicated by **Table 1**, among the respondents with depression, *Dropped interests and activities* was instrumental to measuring the mood symptoms due to the high discrimination value. Among the NMS, *Hallucinations/delusions* had the highest discrimination value. Regarding difficulty, *Double vision* was the most difficult item while the least difficult item was *Preferred to stay at home*.

In **Table 2**, the Wald test was used to compare the individuals with PD to those without PD. Due to the probability of false positives increases due to multiple hypothesis tests, this study primarily reported the adjusted p-values using the Benjamini-Hochberg procedure. Based on this adjustment, six items-*Hallucinations/delusions, Double vision, Unpleasant sensations, Frightening dreams, Dropped interests and activities*, and *Memory difficulties*-were flagged as DIFs. *Excessive sweating* may possibly be flagged as DIF. Compared to the group without PD, the item *Dropped interests and activities* had a difficulty value closer to the median probability of validating this symptom within the group with PD. Meanwhile, this item presented a lower discrimination value for the group with PD. These DIFs may have arisen from bias, so the linear and nonlinear regression analyses were used to evaluate this.

**Table 2.**
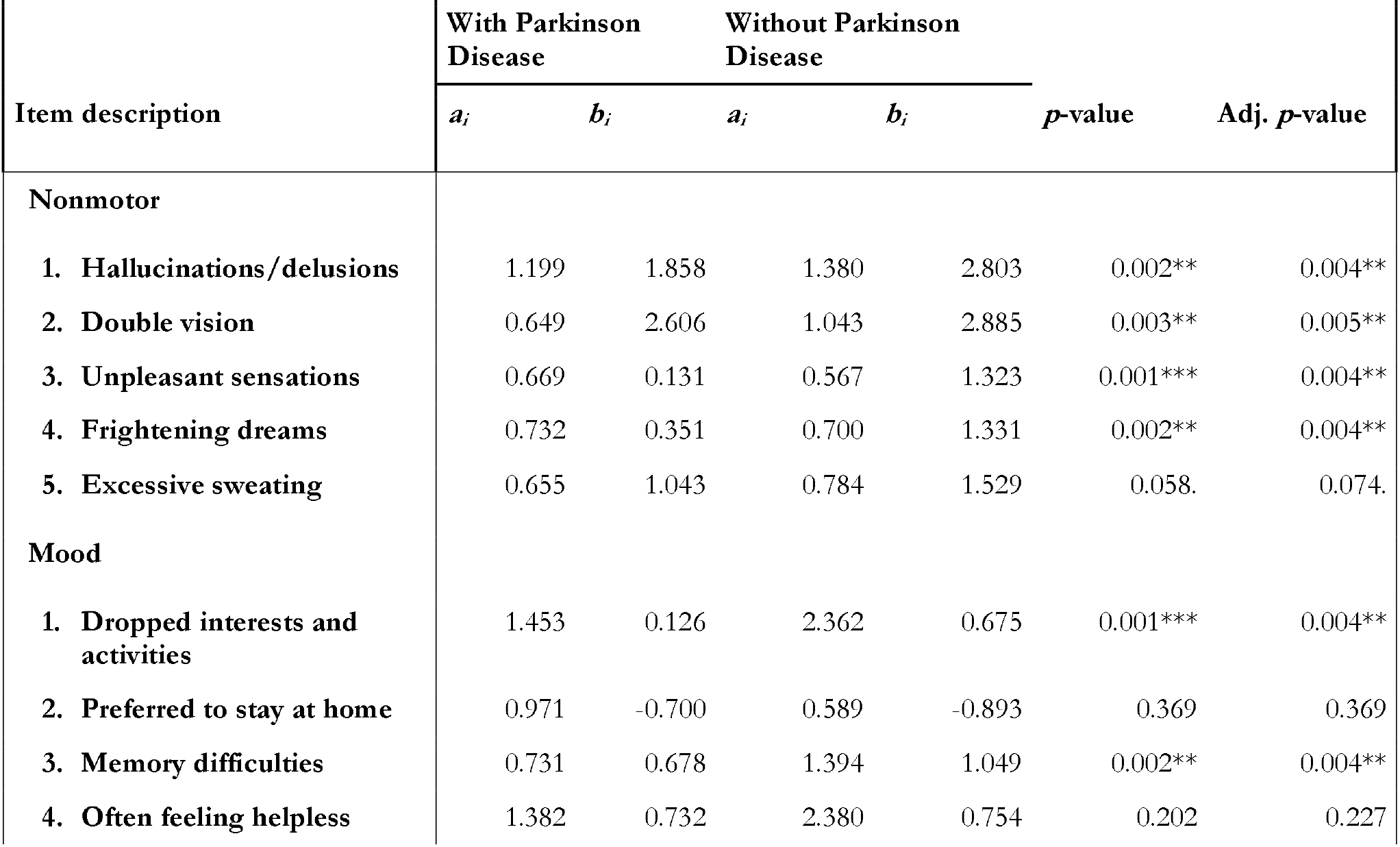
Differential Item Functioning. The significant codes for p-value are 0.001 ‘***’, 0.01 ‘**’, 0.05 ‘*’, and 0.10 ‘.’. The degree of freedom is equal to 2 and the FDR is in use for adjusted *p*-values.

Notably, *Dropped interests and activities* had the highest discrimination value among the depressed individuals both with and without PD (a_i_=1.453, a_i_=2.362). *Unpleasant Sensations* contained the lowest discrimination value among depressed individuals without PD (a_i_=0.567), and *Double vision* resulted in the lowest discrimination value among those with PD (a_i_=0.649). For depressed individuals both with and without PD, *Double vision* was the item with the greatest difficulty (b_i_=2.606, b_i_ = 2.885). *Dropped interests and activities* was least difficult for depressed individuals both with and without PD (b_i_=0.126, b_i_=0.675).

### Regression Model Result

**Table 3** contained two different models with coefficients and two types of standard errors. The dependent variable was the factor score, which was obtained from the expected a posteriori method in the 2-PL model. However, this analysis primarily utilized the Tobit model and bootstrapped standard errors. Due to the presence of the corner solution, the OLS model was not the best linear unbiased estimator. As indicated by the Shapiro-Wilks test, the residuals of both models were not normally distributed. The Fitted vs. Residual plot suggested that the Tobit model was more likely to be homoscedastic. The Breusch-Pagan test determined that the OLS model was homoscedastic as well. However, the maximum likelihood estimation is inconsistent when the residuals are not normally distributed.^22^ The Gaussian parametric bootstrap may remediate this measurement error.

**Table 3.**
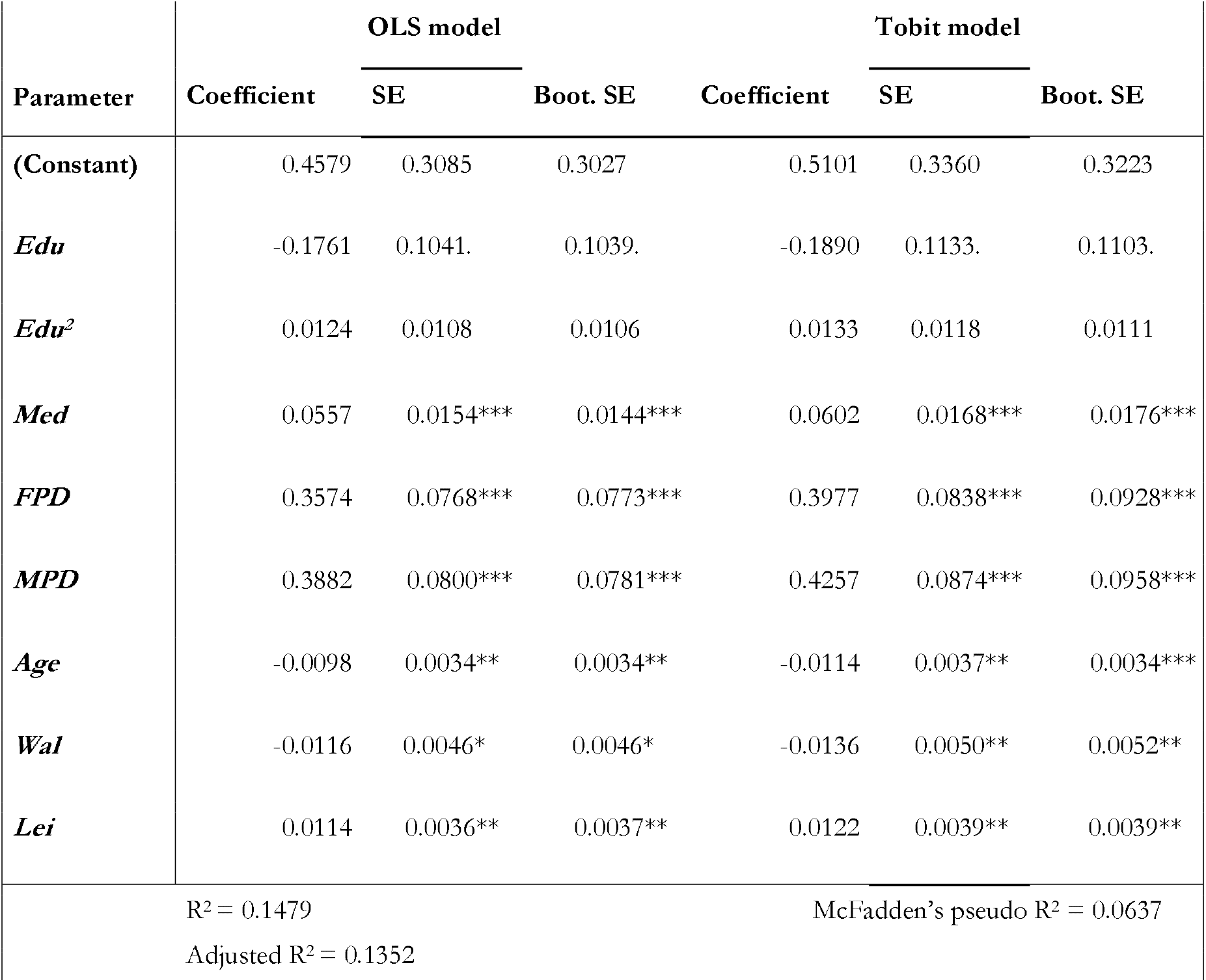
OLS and Tobit Regression Coefficients. The codes for significant p-values are 0.001 ‘***’, 0.01 ‘**’, 0.05 ‘*’, and 0.10 ‘.’ *Edu* is the education attainment. *Med* is a count of unique medicines one has used. *FPD* is a dummy variable for female with PD while *MPD* is a variable for male with PD. *Wal* is average hours of walking per week, while *Lei* is average hours of leisure per week.

| Parameter | OLS model |  |  | Tobit model |  |  |
| --- | --- | --- | --- | --- | --- | --- |
|  | Coefficient | SE | Boot. SE | Coefficient | SE | Boot. SE |
| **(Constant)** | 0.4579 | 0.3085 | 0.3027 | 0.5101 | 0.3360 | 0.3223 |
| ***Edu*** | -0.1761 | 0.1041. | 0.1039. | -0.1890 | 0.1133. | 0.1103. |
| ***Edu*<sup>2</sup>** | 0.0124 | 0.0108 | 0.0106 | 0.0133 | 0.0118 | 0.0111 |
| ***Med*** | 0.0557 | 0.0154*** | 0.0144*** | 0.0602 | 0.0168*** | 0.0176*** |
| ***FPD*** | 0.3574 | 0.0768*** | 0.0773*** | 0.3977 | 0.0838*** | 0.0928*** |
| ***MPD*** | 0.3882 | 0.0800*** | 0.0781*** | 0.4257 | 0.0874*** | 0.0958*** |
| ***Age*** | -0.0098 | 0.0034** | 0.0034** | -0.0114 | 0.0037** | 0.0034*** |
| ***Wal*** | -0.0116 | 0.0046* | 0.0046* | -0.0136 | 0.0050** | 0.0052** |
| ***Lei*** | 0.0114 | 0.0036** | 0.0037** | 0.0122 | 0.0039** | 0.0039** |
| R<sup>2</sup> = 0.1479<br>Adjusted R<sup>2</sup> = 0.1352<br>McFadden's pseudo R<sup>2</sup> = 0.0637 |  |  |  |  |  |  |

Additionally, the Wald-Wolfowitz runs test showed that the residuals of both models were from a random process. The Variance Inflation Factor showed every coefficient was below 2.5 except *Edu*. The added variable *Edu*^*2*^ contributed to its acceptable increase in the Variance Inflation Factor.

The Tobit model had average treatment effects (ATE) equal to 0.3707 and 0.3968 for *FPD* and *MPD*, respectively. The ATE for *Wal* and *Age* was -0.0127 and -0.0098, respectively, while the ATE for *Lei* week was 0.0114. If the ATE was positive, the depression severity was higher. Notably, the ATE for *Med* was 0.0557. The OLS model also had ATE equal to 0.3574 for *FPD* and 0.3882 for *MPD*. Both models concluded that *Edu* had little to no impact on the depression severity in individuals with and without PD.

## DISCUSSION

NMS affects about 30-40% of individuals with PD but most of the treatment options target motor impairments.^3,23^ This is because individuals with PD report motor disabilities more often than NMS.^24^ However, the results from the present study suggest that the presence of both depression and PD-as opposed to depression alone-has a differential impact on the presentation and severity of NMS.

Tethered with our hypothesis, *Dropped interests and activities* has a notably strong negative impact on both populations in this study, especially on the depressed individuals with PD. *Dropped interests and activities* was most severe within both populations. This behavior-changing psychological symptom most commonly stems from apathy and is consistent with current literature.^25^ Apathy often co-occurs with depression among individuals with PD, and depression is one of the most common NMS that these affected individuals experience.^26^ Apathy can also influence medication use, as this additional symptom to PD can warrant additional treatment methods.^27,28^ This association is in line with our findings. We have found an increasing severity of the apathetic symptom *Dropped interests and activities*, but the actual prevalence of this symptom is most likely much higher. Considering these findings, it is important that both PD patients and professionals are aware of the relationships between PD, depression, and apathy for early potential identification of associated symptoms and treatment.

Even though one sensory symptom-*Double vision*-was not highly associated with the population of individuals with and without PD, the DIF was flagged between these groups. This phenomenon should further be studied.

**Table 3** indicates that while holding other variables constant, the increase in *Med* leads to the increase in factor score at the 1% significance level; medication use, especially use of numerous medications, can cause a variety of side effects, potentially exacerbating NMS. While the results show that greater average number of hours walking was associated with decreased depression severity, the increased average number of leisure hours was associated with greater depression severity. Exercise is a common recommendation to alleviate depressive symptoms, and these results are mirrored for individuals with both depression and PD. Exercise had a significant, positive influence on the co-occurring diseases. As an individual’s depression worsens, there is an overall upward trend in the severity of their NMS.^29^ Thus, particularly when depression is also present, the treatment of both depression and NMS is crucial alongside motor symptom-focused treatment options to improve the affected individual’s QOL.^24^

The large overall sample size of respondents from Fox Insight is a clear strength of this study-allowing for greater reliability and credibility. Another strength lies in the utilization of latent variable analysis to treat each item differently and to apply model specification for linear and nonlinear regressions with a theoretical framework in mind. Undertaking this latent variable and causal analysis to explore specific symptom differences among depressed individuals with and without PD contributes to general knowledge by identifying population-specific support needs.

Despite the high concurrence rate between self-report and clinician-determined diagnoses, this database remains susceptible to biases and errors due to the sensitive nature of some questions.^30^ Another limitation of this study was that the dataset was primarily PD-focused, resulting in small sample sizes for individuals with depression compared to the overall respondents in the Fox Insight study. Additionally, the generalizability is limited due to the underrepresentation of minority populations and education levels. The latent variable analysis was subject to an item process that became susceptible to overfitting, as indicated by the infit standardized mean statistics. Furthermore, the dependent variable was based on predicted values rather than observed values, and non-normal residuals also weakened the Tobit model. Finally, the manual approach with assumptions used in constructing the directed graph needs to be reevaluated using different methods, especially considering the larger sample size.

## CONCLUSION

From this study, we recommend specific steps for future research studies to further address the research questions. For example, a thematic analysis should be conducted to investigate the surprising finding regarding the relationship between the male group with PD and severity of depressive symptoms. Additionally, to assess the generalizability of latent variable analysis model, this model needs to be reevaluated using a new dataset. With a larger dataset, two causal discovery algorithms, fast greedy equivalence search, and fast causal inference should be employed to evaluate the identification and consistency of this directed graph. Furthermore, apathy and depression are often reported together, posing challenges to determine whether depression is a latent variable. Apathy can mimic depression, which makes apathy difficult to address as well. The specific effects of depression and apathy on NMS should also be further explored.

## Data Availability

All data produced in the present study are available upon reasonable request to the authors.
All data produced in the present work are contained in the manuscript.
All data produced are available online at FoxDEN Insight (https://foxden.michaeljfox.org/insight/explore/insight.jsp).

## ACKNOWLEDGEMENTS

The authors thank Dr. Monica Javidnia for the initial mentorship and support given for this project. The authors also thank the University of Rochester, the Rochester Institute of Technology, and Gallaudet University for providing the resources and programs used to bring this research initiative to fruition.

## ABOUT THE STUDENT AUTHOR

Jasmine Moon graduated from the University of Rochester in May 2022.

## PRESS SUMMARY

Nonmotor symptoms occur at the early stages of Parkinson’s Disease and can have a greater impact on an individual’s quality of life compared to motor symptoms. Additionally, at least 50% of people with Parkinson’s Disease experience depression, but the impact of the severity of nonmotor symptoms due to the comorbidity of these two diseases is unclear. This study aims to analyze and compare the magnitudes of nonmotor symptoms in depressed individuals with and without Parkinson’s Disease. Data is extracted from the longitudinal study Fox Insight, and statistical analysis is performed through the directed acyclic graph approach and the item response theory.

